# Quetiapine in a Low Dosage During Pregnancy: Neonatal Outcomes in a Retrospective Cohort Study

**DOI:** 10.64898/2026.01.18.26344354

**Authors:** Sydney FJ van Tienoven, Flora Gossink, Remke C Dullemond, Jeroen van Dillen

## Abstract

**Background:** Prescribing pharmaceutical agents during pregnancy requires a balance between maternal therapeutic needs and foetal risks. Quetiapine, an atypical antipsychotic, is prescribed in psychiatric disorders and increasingly used off-label in low-dosages (<200mg daily) for insomnia during pregnancy. This study aimed to investigate the possible relationship between using quetiapine in a low dosage during pregnancy and the neonatal outcome within the first three days after birth.

**Methods:** A retrospective cohort study was conducted at the Radboud University Medical Centre (RadboudUMC), Nijmegen, The Netherlands. Liveborn neonates admitted to the RadboudUMC during the years 2017-2024 whose mothers used at least one dose of quetiapine (≤ 200mg daily) during pregnancy were included. Maternal characteristics and neonatal outcomes were analysed using SPSS, dose-dependent outcomes were also assessed.

**Results:** 30 neonates were included. Of these, 7 (23.3%) were born preterm, 7 (23.3%) were small for gestational age (SGA), 6 (20.0%) were admitted to the neonatal intensive care unit (NICU) and 2 (6.3%) exhibited withdrawal symptoms. Most neonates were admitted to the hospital for 3 days.

**Conclusion:** This study is the first to evaluate neonatal outcome after maternal low dose quetiapine in pregnancy in the Netherlands. We found relatively high rates of preterm birth, SGA and NICU admissions. As this was a single-centre study without a control group and with a small sample size in an academic population, results should be interpreted with caution. We recommend further prospective and multicentre studies including control groups.

## Introduction

The use of pharmaceutical agents during pregnancy presents a complex clinical challenge, requiring a careful balance between therapeutic needs of the mother and potential risks to the developing foetus. Among medicines prescribed for various psychiatric disorders during gestation, atypical antipsychotics have received significant attention due to their potential impact on maternal and neonatal health outcomes^1^.

Quetiapine, an atypical antipsychotic drug, is used in the treatment of conditions such as schizophrenia, bipolar disorder, and severe depressive disorder^2^. Quetiapine differs from other atypical antipsychotics in its dose-dependent effect. Low doses (25-100mg daily) show sedative effects, doses of 300-600mg daily result in mood stabilization and anxiety reduction and high doses (> 800mg daily) are used in schizophrenia^2^. It is increasingly used off-label in the treatment of agitation and posttraumatic stress disorder (PTSD). A recent study showed efficacy of a low dosage quetiapine in insomnia^3^. Common side effects are extrapyramidal symptoms at high dosages and metabolic risks including weight gain and hypertension^4,5^.

Most studies on quetiapine during pregnancy have focused on high dosage regimens. Some studies showed significantly adverse neonatal outcomes such as lower Apgar scores, extrapyramidal disorders, and neonatal withdrawal symptoms, particularly when quetiapine was administered during the third trimester^6,7^. Another study showed an increased risk of developing gestational diabetes, which is associated with neonatal hypoglycaemia and macrosomia^8^. One study states an increased risk for low birth weight in neonates with prenatal exposure to quetiapine^9^. Yet, psychiatric disorders themselves also increase the risk of complications such as preterm birth and low birth weight^10,11^.

The use of quetiapine during the first and second trimesters is considered probably safe and is not associated with a higher risk of developing major congenital malformations^12^.

Despite this, usage of low-dose quetiapine during pregnancy has not been investigated before. An increased understanding of the effect of low-dose quetiapine on neonatal outcomes will help in future clinical decision-making. Therefore, this study aims to investigate the relationship between prenatal exposure to low-dose quetiapine and neonatal outcomes within the first three days after birth.

## Methods

### Study Design and Setting

A retrospective single-centre cohort study was conducted at the Radboud University Medical Centre (RadboudUMC) in Nijmegen, The Netherlands. A tertiary academic medical centre which manages around 1,500 deliveries annually^13,14^. The centre includes a Psychiatric, Obstetric, and Paediatric (POP) department where a multidisciplinary approach is used to care for obstetric patients with psychiatric problems, considering the possible impact on the unborn foetus. Approximately one in ten women experience psychiatric complaints during pregnancy or postpartum^15^.

### Inclusion and Exclusion Criteria

Figure 1 shows the flowchart of patient inclusion and exclusion. The study included liveborn neonates admitted to the RadboudUMC between January 1^st^, 2017, and May 1^st^, 2024, whose mothers used at least one dose of quetiapine during pregnancy (first, second and/or third trimester) in a low dosage (≤ 200mg). Excluded were stillborn neonates and those whose parents declined participation through the ‘no objection procedure’. A list of possible subjects was generated from the electronic patient file system (EPIC) with the terms ‘pregnancy’ and ‘quetiapine’.

**Figure 1.**
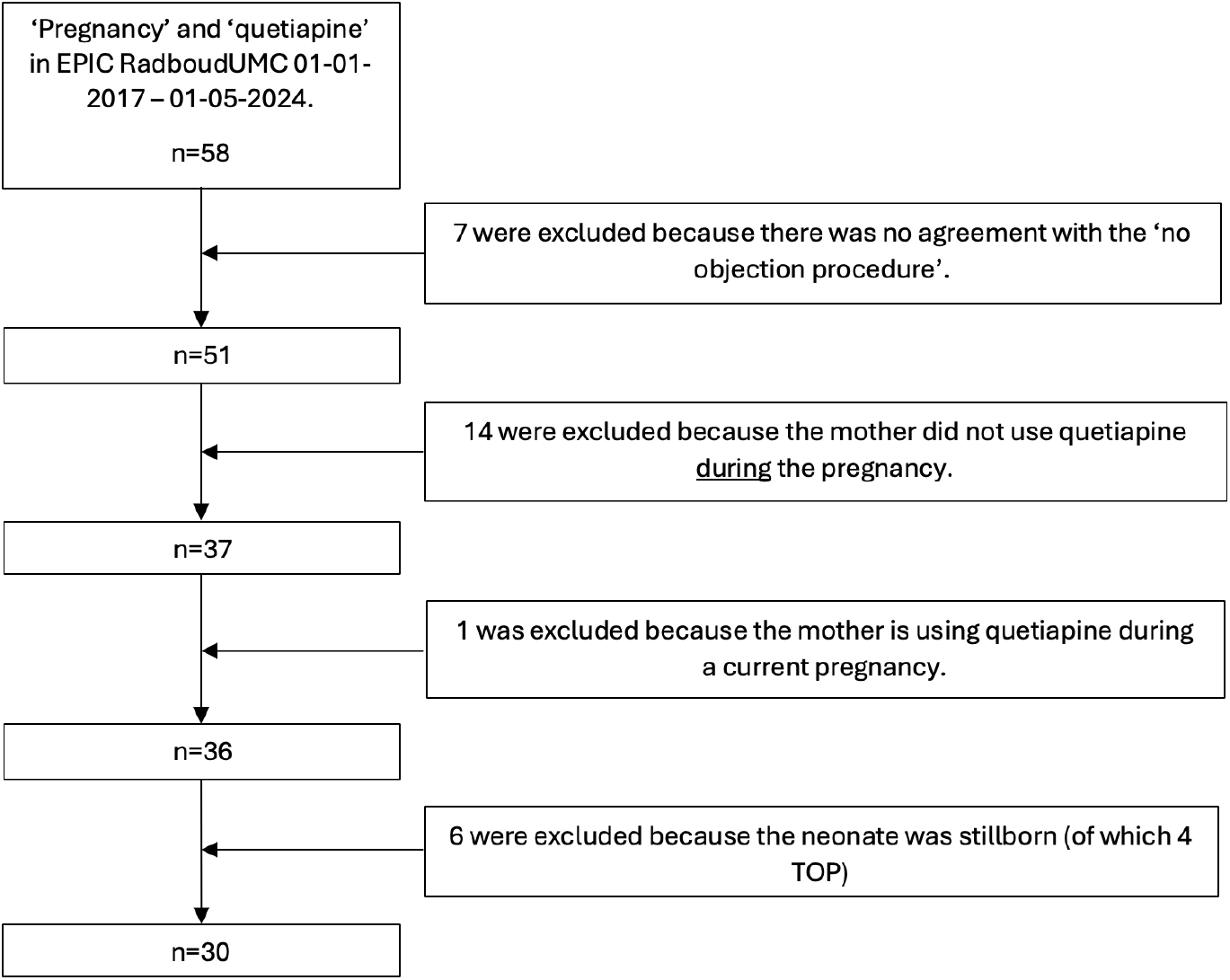
Flowchart patient in- and exclusion

Six cases were excluded due to stillbirth, four of which were terminations of pregnancy (TOP) due to various congenital defects.

### Variables and Measurements

Maternal characteristics included ethnicity, date of birth, gravidity, parity, body mass index (BMI) at the beginning of pregnancy, indication for and dosage of quetiapine, usage during third trimester, other psychotropic and overall medication usage, intoxications and mode of delivery. Maternal age at childbirth was calculated. If BMI was not stated, it was calculated using the ‘Voedingscentrum’ calculator with recorded weight and height and categorized as underweight (<18.5), healthy weight (18.5-25), overweight (25-30), and obese (>30)^16^. As amphetamines are often prescribed to psychiatric patients and are associated with possible neonatal adverse events, including withdrawal symptoms, this medication was specifically noted^17^.

Neonatal outcomes focused on preterm birth (gestational age < 37 weeks^18^), asphyxia, withdrawal symptoms, birth weight, head circumference and NICU admission (including length of stay and interventions). Asphyxia was diagnosed if cord pH <7.0, BE <-16 mmol, and Apgar score <5 at 5 minutes^19^. Birth weight was evaluated using the Hoftiezer curve adjusted for gestational age and gender, categorizing neonates as small for gestational age (SGA, p < 10) or large for gestational age (LGA, p > 90); macrosomia was defined as a birth weight ≥ 4500 gram as stated in the NVOG guideline^20-22^. Finnegan scores were used to quantify withdrawal symptoms for neonatal abstinence syndrome (NAS)^23^.

These outcomes were compared with data from Perined and Peristat representing the Dutch population in 2021, including birth weight and NICU admission rates among term neonates, and incidences of low Apgar scores (<7) and preterm births across the population^24,25^.

### Statistical Analysis

Statistical analysis was performed using SPSS. The Shapiro-Wilk test assessed the distribution of numeric variables. Normally distributed variables (maternal age, BMI, quetiapine dosage, birth weight, cord pH, cord BE) were analysed with means and standard deviations. Non-normally distributed variables (gravidity, parity, gestational age, head circumference, length of hospital stay, Apgar score) were analysed with medians and confidence intervals. Multi-answer variables were used for interventions and paediatric evaluations. Data was coded and entered into SPSS for analysis. Descriptive frequency tables were generated for maternal characteristics and neonatal characteristics and outcomes. A chi-square test was conducted to examine dose-dependency of primary neonatal outcomes across quetiapine dosage categories (<25mg, 25-100mg, >100mg). National data from Perined and Peristat was used for comparison^24,25^. As some data applied to term neonates, in SPSS a filter was applied to isolate term birth data, allowing for comparative analysis.

### Ethical Considerations

The study protocol was reviewed and declared not needing ethical approval under the Dutch Medical Research Involving Human Subjects Act (WMO) by the Medical Ethics Committee Oost-Nederland (METC-Oost) (file number: 2024-17255). Data collection was facilitated through the ‘no objection procedure’ of the RadboudUMC. This procedure implies that patients consent for their data use for research purposes unless they explicitly opt out.

All collected data will be securely stored in the RadboudUMC system for a period of 15 years.

## Results

Between January 1^st^, 2017, and May 1^st^, 2024, 58 neonates born to mothers who used quetiapine were admitted to the RadboudUMC. After applying exclusion criteria, 30 neonates were included.

### Maternal Characteristics

Maternal characteristics are presented in Table 1. Most mothers were Caucasian (n=28, 93.3%). All with psychiatric histories, including depression, social anxiety disorder, autism, post-traumatic stress disorder (PTSD), or borderline personality disorder, which were the primary indications for quetiapine. In four patients (13.3%) insomnia was the indication, with dosages between 6.25mg and 25mg daily. Higher doses were used for psychotic symptoms (175mg) and PTSD (150mg). Most cases (n=21, 70.0%) involved dosages from 25 to 100mg, the majority continued during the last trimester of pregnancy (n=28, 93,3%).

**Table 1.**
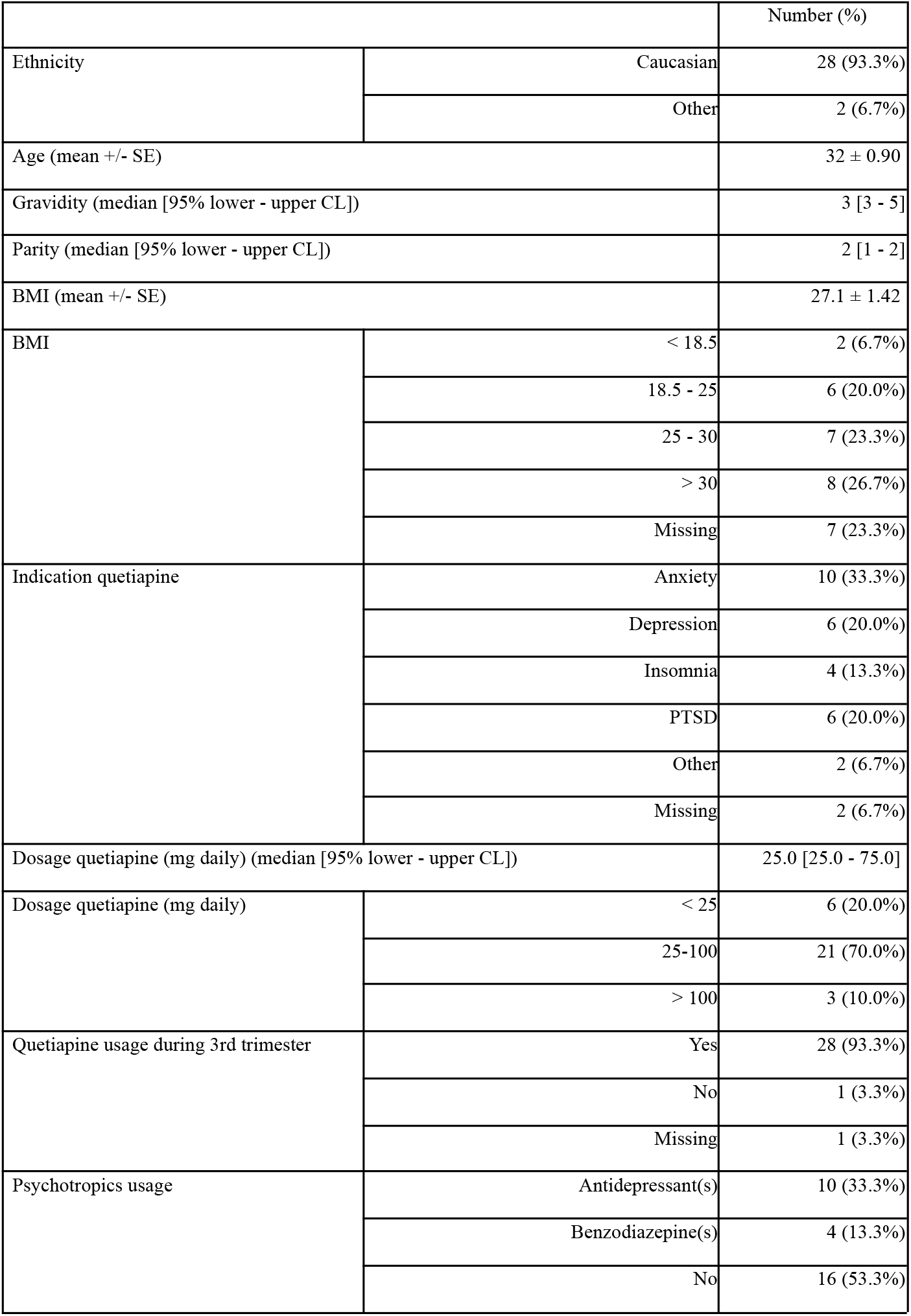
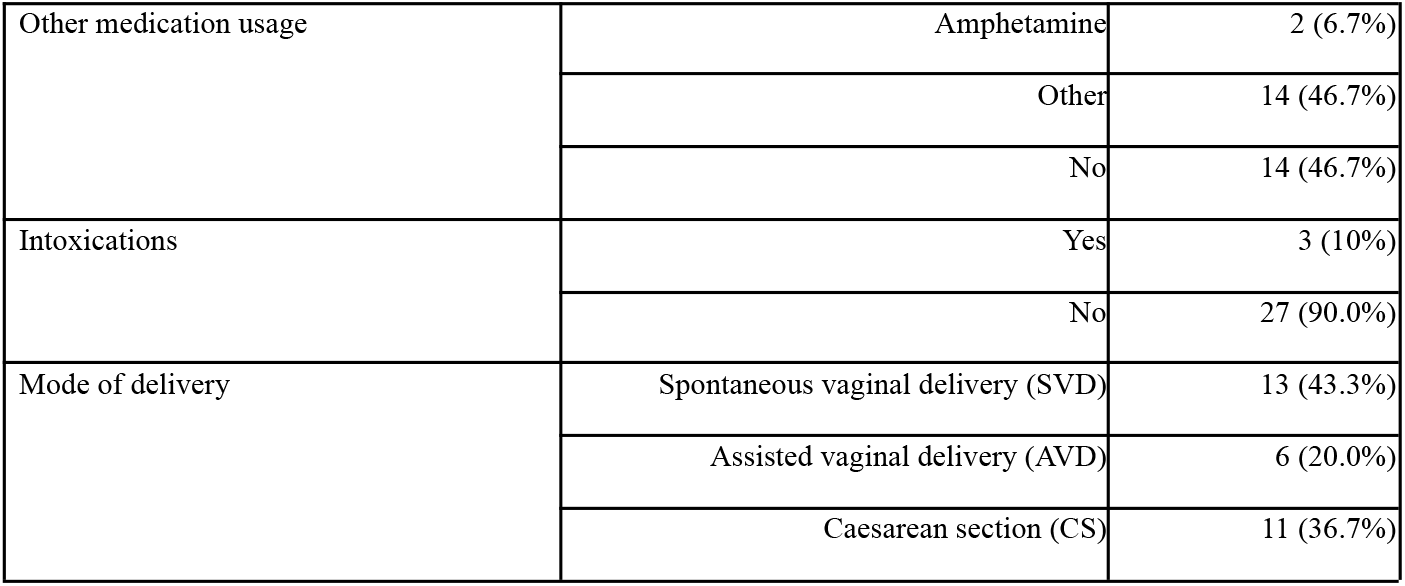
Maternal characteristics during pregnancy of study population.

The mean BMI at the beginning of pregnancy was 27.1; with 23.3% classified overweight (n=7) and 26.7% obese (n=8). Related obstetric conditions included gestational hypertension (n=6, 20.0%) and gestational diabetes (n=5, 16.7%). Half of the mothers with hypertension (n=3) and 60.0% of those with diabetes (n=3) required medication. Two mothers used amphetamines, ten antidepressants (33.3%), and four benzodiazepines (13.3%). Over half of the mothers (53.3%) did not use other psychotropic medications.

### Neonatal Characteristics and Outcomes

Table 2 presents neonatal characteristics and outcomes. Table 3 shows outcomes per dosage. Figure 2 illustrates a comparison of outcomes with data from national data^24,25^.

**Table 2.**
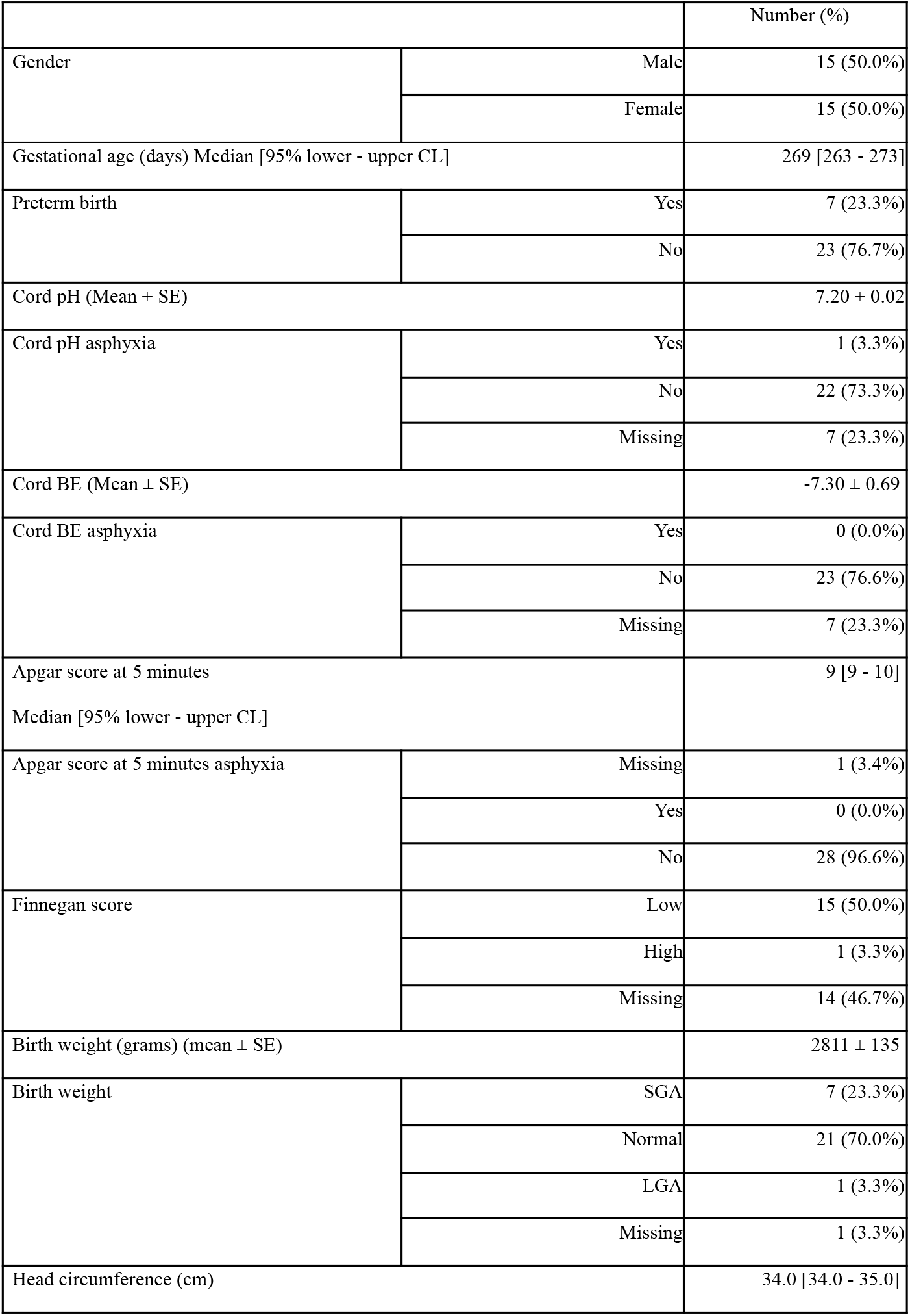
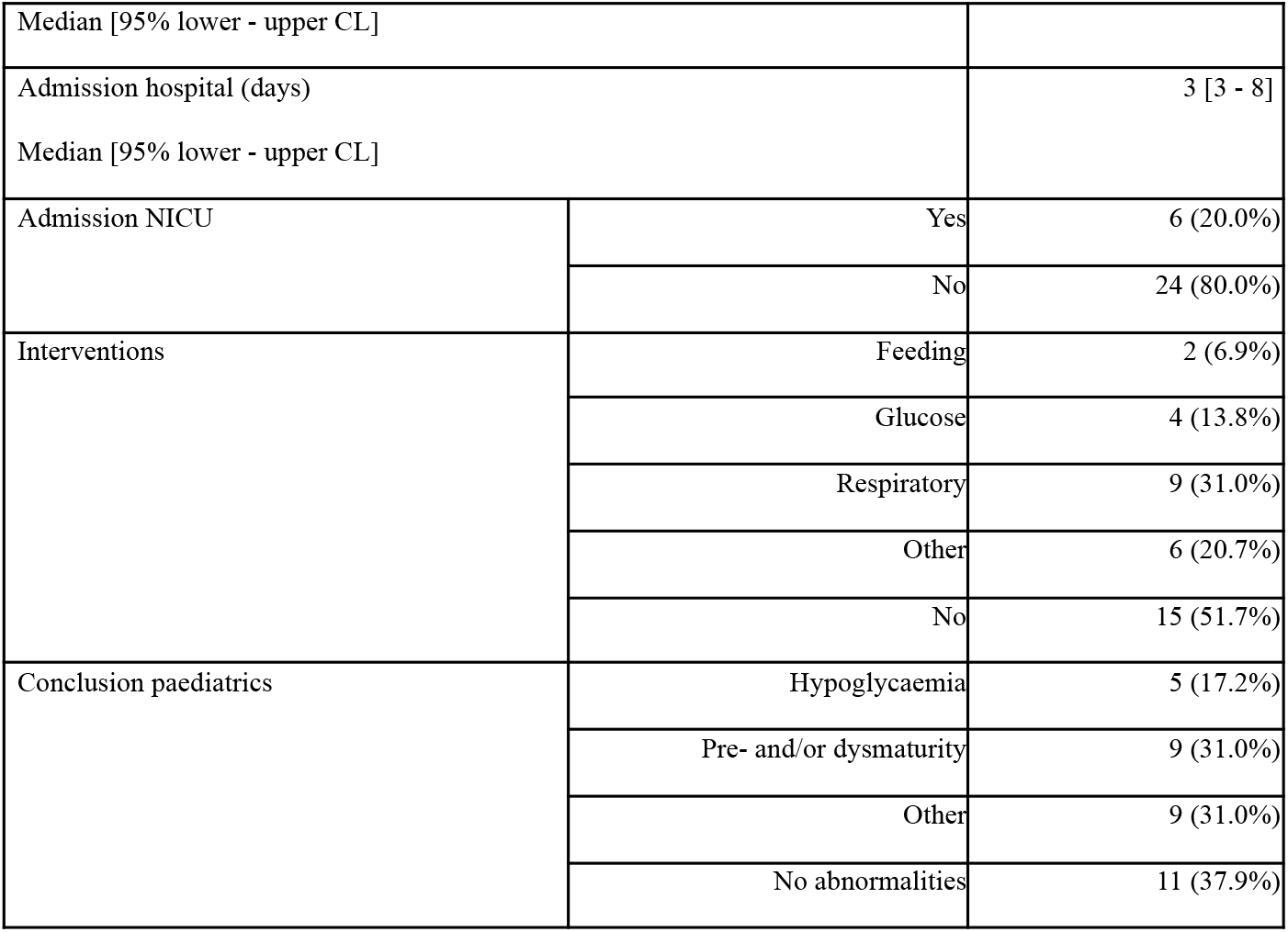
Neonatal characteristics and outcomes.

**Table 3.** Neonatal primary outcome measurements per subgroup of dosage quetiapine.

|  |  | Dosage quetiapine<br>(mg daily) |  |  | Total | Value Pearson Chi-square | df | Asymptotic significance |
| --- | --- | --- | --- | --- | --- | --- | --- | --- |
|  |  | < 25 | 25-100 | > 100 |  |  |  |  |
| Preterm<br>birth | Yes | 3 | 3 | 1 | 7 |  |  |  |
|  | No | 3 | 18 | 2 | 23 |  |  |  |
| Total |  | 6 | 21 | 3 | 30 | 3.541 | 2 | 0.173 |
| Birth weight | SGA | 1 | 5 | 1 | 7 |  |  |  |
|  | Normal | 5 | 14 | 2 | 21 |  |  |  |
|  | LGA | 0 | 1 | 0 | 1 |  |  |  |
|  | Missing | 0 | 1 | 0 | 1 |  |  |  |
| Total |  | 6 | 21 | 3 | 30 | 1.293 | 6 | 0.972 |
| Admission<br>to NICU | Yes | 2 | 3 | 1 | 6 |  |  |  |
|  | No | 4 | 18 | 2 | 24 |  |  |  |
| Total |  | 6 | 21 | 3 | 30 | 1.429 | 2 | 0.490 |

**Figure 2.**
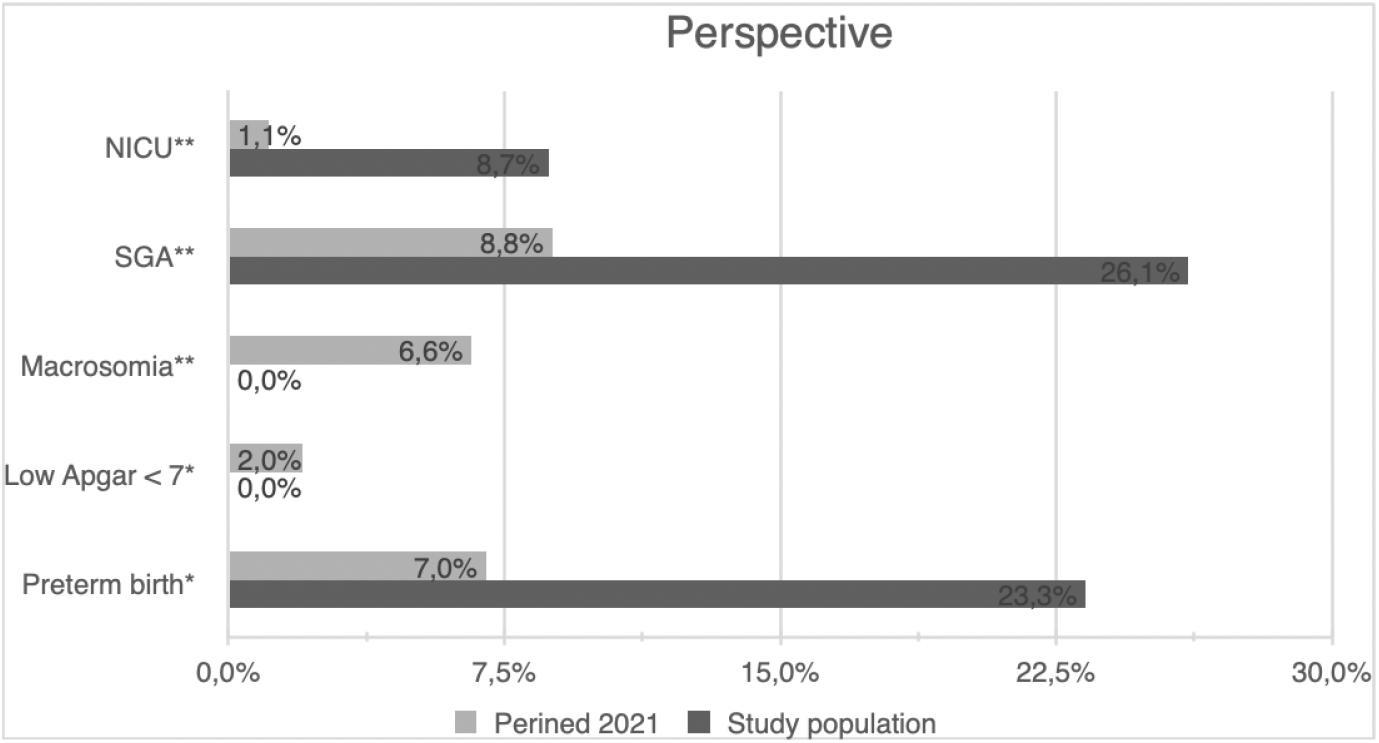
Neonatal primary outcomes of the study population compared to the general population in the Netherlands in the year 2021 (obtained from Perined and Peristat (24, 25)) ^*^All neonates ^**^Term neonates

#### Preterm Birth

Among the neonates in our study, 23.3% (n=7) were born preterm. Two (6.7%) occurred in a twin pregnancy delivered via caesarean section (CS) due to complicated maternal COVID-19 infection. Two neonates (6.7%) were born preterm due to premature rupture of membranes (PROM). The youngest, born at 28 weeks via CS due to foetal growth restriction (FGR) and abnormal Dopplers, was the only neonate classified as SGA (p<3) in this subgroup, receiving iron suppletion for anaemia. Three preterm neonates (10.0%) received respiratory support via continuous positive airway pressure (CPAP).

There was no significant difference across the dosage subgroups of <25mg, 25-100mg, and >100mg (p=0.173).

#### Asphyxia

Among the neonates in our study, none showed signs of asphyxia based on cord blood BE or Apgar score. One neonate (3.4%) exhibited asphyxia based on a cord pH of 6.99. Despite this, the neonate had a reassuring Apgar score of 9/10/10. This neonate was born at term and mother used 50mg quetiapine without other psychotropic medications. The reason for the low cord pH remains unclear.

There were no significant differences between the dosage subgroups in cord pH (p=0.786), cord BE (p=0.535), or Apgar score (p=0.535).

#### Withdrawal Symptoms

Among the neonates for whom the Finnegan score was reported (n=16), 15 (93.8%) had a low score, while one (6.3%) had a high score. The neonate with the high score was born at term via spontaneous vaginal delivery (SVD) to a mother using 100mg quetiapine and sertraline, with no other medications or intoxications. This neonate showed no other abnormal primary outcomes but required feeding intervention for neonatal hypoglycaemia. Mother had a BMI of 27.5 and no history of gestational diabetes.

There was no significant difference in Finnegan scores across the subgroups (p=0.837)

#### Birth Weight

The birth weight of the majority of neonates in our study fell between the 11th and 50th percentiles (n=14, 48.3%). One neonate (3.3%) was classified LGA with a Hoftiezer p-value >95, but not macrosomic, with a birth weight of 4002 grams. This neonate was born at term following induction of labour due to suspected macrosomia and did not exhibit neonatal hypoglycaemia. Mother was on 100mg quetiapine and 10mg amitriptyline daily and had gestational diabetes managed through diet, with a BMI of 25.73.

Seven neonates (23.3%) were classified SGA. Among the term births, 26.1% was classified SGA. Notably, 57.1% of mothers in the SGA subgroup also used other psychotropic medications. Two neonates were suspected of having a syndrome, of which one was present in the mother and one was prenatally exposed to smoking. One neonate in the SGA group required CPAP.

No significant difference in birth weights was seen across the subgroups (p=0.865).

#### NICU Admission

The rate of NICU admissions among term neonates in our study population was 8.7%. Most neonates were admitted to the hospital for three days. The longest admission lasted 33 days due to prematurity and dysmaturity in a twin pregnancy. Other extended stays lasted 18 and 8 days, also due to prematurity and dysmaturity. Six neonates (20.0%) were admitted to the NICU: four for prematurity and/or dysmaturity, one for respiratory distress, and one for the development of persistent pulmonary hypertension of the newborn (PPHN).

There was no significant difference in NICU admissions among the subgroups (p=0.490).

Interventions administered to neonates included feeding (n=2, 6.9%), glucose (n=4, 13.8%), respiratory support (n=9, 31.0%), and other treatments (n=6, 20.7%), such as phototherapy (n=2) and antibiotics (n=3). Paediatricians concluded that 11 neonates (37.9%) had no abnormalities, nine (31.0%) were premature and/or dysmature, three (10.3%) developed hypoglycaemia, and nine (31.0%) had other conditions.

## Discussion

This study provides insights into neonatal outcomes associated with maternal quetiapine use in low dosages during pregnancy, revealing a relatively high rate preterm birth (23.3%), NICU admission (8.7%) and SGA (26.1%). No cases of asphyxia, low Apgar score or macrosomia were observed. One neonate exhibited withdrawal symptoms. The observed higher prevalence of preterm births is noteworthy, considering previous studies have not found this association^7,26^. However, another study showed an increased risk of preterm birth among mothers with psychiatric disorders, which could be a confounding factor in our study population^11^.

The increased prevalence of SGA is important, as prior research has not established a link between quetiapine use and SGA^27,28^. This elevated rate may have been influenced by logistic factors as the single centre academic population or medical factors as mother’s psychiatric condition and use of other psychotropic medications^10^.

Interestingly, our study did not find cases of macrosomia, despite previous associations between quetiapine use, gestational diabetes and macrosomia^8^.

In contrast to prior research that reported lower Apgar scores in neonates exposed to high dosages, our study found no decrease in Apgar scores at 5 minutes^7,28^. This may be attributed to the lower dosage of quetiapine used in our study.

Only one neonate exhibited withdrawal symptoms, which contrasts with a prior study that reported a significant higher amount of withdrawal symptoms in neonates prenatally exposed to quetiapine^28^. This may be because in our study a lower dosage was used than in the previous study^28^. However, the current Dutch guideline for maternal atypical antipsychotic use recommends monitoring new-borns for at least 24 hours for withdrawal symptoms^29^.

A strength of our study is that it examines quetiapine in a low dosage during pregnancy, an area that has not been investigated before. This allows exploration of neonatal outcomes associated with low dosages, providing insight into its safety in pregnancy. However, several limitations need to be acknowledged. Therefore, the findings of our study need to be interpreted with caution.

An important limitation is the small sample size of our study population and the absence of a control group. These factors limit the generalizability of our findings and increase the potential for bias. Uneven subgroup sizes make it challenging to clearly see differences in outcomes between the dosages.

Another limitation is the retrospective design, which may have caused bias through confounding factors like the use of other psychotropic medications.

Additionally, conducting the study in a tertiary care facility could have influenced our findings. Patients with threatened preterm births (< 32 weeks) or complicated congenital malformations are likely to be referred to third-line healthcare facilities, potentially explaining the higher rate of preterm births and congenital malformations observed in our population. Also, tertiary centres tend to manage more complex obstetric cases, which may require higher rates of NICU admissions. Therefore, our study population may not be representative to the general pregnant population.

Bias could also be attributed to the inclusion of twins and neonates from the same mother (five times). Furthermore, missing data on important variables, such as the Finnegan score and maternal BMI, limit our ability to draw conclusions.

Some psychiatrists prefer prescribing benzodiazepines for conditions such as insomnia instead of quetiapine, aiming to avoid the potential risk of metabolic syndrome. However, benzodiazepine use during pregnancy has also been associated with adverse outcomes^31^. Showing an increased risk of preterm birth, low birth weight, smaller head circumferences, low Apgar score, NICU admissions, respiratory distress syndrome and withdrawal symptoms^32,33^. Additionally, the ‘floppy infant syndrome’ is reported in literature, characterized by hypotonia, lethargy, disordered temperature regulation, and poor feeding^34^.

It is important to consider that these findings may be influenced by confounding factors such as psychiatric symptoms^31^. If future studies show safety of quetiapine this could be a viable alternative for benzodiazepines for treating insomnia, considering the comparative risks of both pharmaceutical agents for both mother and neonate.

A prospective study with a larger sample size and a control group is recommended to better understand the possible relationship between quetiapine use in a low dosage during pregnancy and adverse neonatal outcomes. A multicentre study, including data from secondary healthcare facilities, could help reduce selection bias. Also, investigating the association between quetiapine use, gestational diabetes and neonatal outcomes as macrosomia, is advised to explore its metabolic consequences during pregnancy.

## Data Availability

All collected data will be securely stored in the RadboudUMC system for a period of 15 years.

